# Dissecting the association of C-reactive protein levels with PTSD, traumatic events, and social support

**DOI:** 10.1101/19009134

**Authors:** Carolina Muniz Carvalho, Frank R Wendt, Adam X. Maihofer, Dan J. Stein, Murray B. Stein, Jennifer A. Sumner, Sian M. J. Hemmings, Caroline M. Nievergelt, Karestan C. Koenen, Joel Gelernter, Sintia I Belangero, Renato Polimanti

**Affiliations:** Department of Psychiatry, Yale School of Medicine and VA CT Healthcare Center, West Haven, CT 06516, USA; Department of Psychiatry, Universidade Federal de São Paulo (UNIFESP), São Paulo, SP, Brazil; Genetics Division, Department of Morphology and Genetics, Universidade Federal de São Paulo (UNIFESP), São Paulo, SP, Brazil; Department of Psychiatry, School of Medicine, University of California, San Diego, La Jolla, CA, USA; Center of Excellence for Stress and Mental Health, Veterans Affairs San Diego Healthcare System, San Diego, CA, USA; MRC Unit on Risk & Resilience in Mental Disorders, Department of Psychiatry and Neuroscience Institute, University of Cape Town, Cape Town, South Africa; Psychiatry Service, Veterans Affairs San Diego Healthcare System, San Diego, CA, USA; Department of Psychology, University of California, Los Angeles, Los Angeles, CA, USA; Department of Psychiatry, Faculty of Medicine and Health Sciences, Stellenbosch University, Cape Town, South Africa; Department of Epidemiology, Harvard School of Public Health, Boston, MA, United States; Departments of Genetics and Neuroscience, Yale University School of Medicine, New Haven, CT 06510, USA

## Abstract

Inflammatory markers like C-reactive protein (CRP) have been associated with posttraumatic stress disorder (PTSD) and traumatic experience, but the underlying mechanisms are unclear. We investigated the association among CRP, PTSD, and traits related to traumatic events and social support using genome-wide data from the Psychiatric Genomics Consortium (30,000 cases and 170,000 controls), the UK Biobank (UKB; up to 117,900 individuals), and the CHARGE study (Cohorts for Heart and Aging Research in Genomic Epidemiology, 148,164 individual). Linkage disequilibrium score regression, polygenic risk scoring, and two-sample Mendelian randomization analyses were used to investigate genetic overlap and causal relationships. Genetic correlations of CRP were observed with PTSD (rg=0.16, p=0.026) and behavioral and emotional response to trauma, exposure to traumatic events, and the presence of social support (−0.28<rg<0.20; p<0.008). We observed a bidirectional association between CRP and PTSD (CRP→PTSD: β=0.065, p=0.015; PTSD→CRP: β=0.008, p=0.009). CRP also showed a negative association on the “felt loved as a child” trait (UKB, β=-0.017, p=0.008). Due to the known association of socioeconomic status (SES) on PTSD and social support, a multivariable MR was performed to investigate SES as potential mediator. We found that household income (univariate MR: β=-0.22, p=1.57×10^−7^; multivariate MR: β=-0.17, p=0.005) and deprivation index (univariate MR: β=0.38, p=1.63×10^−9^; multivariate MR: β=0.27, p=0.016) were driving the causal estimates of “felt loved as a child” and CRP on PTSD. The present findings highlight a bidirectional association between PTSD and CRP levels, also suggesting a potential role of SES in the interplay between childhood support and inflammatory processes with respect to PTSD risk.

## Introduction

Posttraumatic stress disorder (PTSD) is a common psychiatric condition that can occur after experiencing or witnessing traumatic events.^1^ Inflammatory processes, reflected by blood-based markers like C-reactive protein (CRP), could be involved in the biology of the trauma response and PTSD.^2^ CRP is a protein that responds to inflammatory stimuli by triggering cellular reactions such as those related to macrophage recruitment during inflammation initiation.^3^ Elevated levels of pro-inflammatory cytokines (e.g. interleukins; a downstream product of CRP signaling) and acute-phase proteins (e.g. CRP) have been observed in the blood of individuals with PTSD.^4, 5^ Meta-analyses of cross-sectional studies have confirmed the association of inflammation with traumatic events and PTSD.^6,7^ Longitudinal studies reported evidence supporting a bidirectional association: elevated inflammation may contribute to PTSD and that PTSD contributes to elevated inflammation.^8-13^ Additionally, PTSD is well-known to be associated with many chronic diseases (e.g., cardiovascular diseases, type-2 diabetes, and rheumatoid arthritis) with an established inflammatory component.^14-16^ Understanding the mechanisms underlying the PTSD-inflammation association can potentially lead to important translational implications. If inflammation is causal for PTSD, inflammatory markers may be useful in identifying persons at risk, and anti-inflammatory treatments may be therapeutic. If PTSD causes inflammation, this would provide evidence for one pathway via which PTSD increases risk of chronic disease and may support screening for inflammation in persons with PTSD. A third hypothesis is that the relation between inflammation and PTSD is not causal in either direction but accounted for by a third factor.

Analytic approaches based on large-scale genome-wide datasets may help us to understand how inflammatory processes and CRP levels are related to etiology or consequences of PTSD. Mendelian randomization (MR) can be used to investigate causal relationships using genetic variants (e.g., single nucleotide polymorphisms – SNPs) as instrumental variables in genetic epidemiologic experiments analogous to randomized control trials.^17-19^ In this context, we investigated the genetic overlap and the putative causal relationships linking CRP and PTSD, leveraging large-scale genome-wide datasets from the Psychiatric Genomic Consortium PTSD Workgroup (PGC-PTSD), the UK Biobank (UKB), and the Cohorts for Heart and Aging Research in Genomic Epidemiology (CHARGE) study. To investigate more broadly the relationship between trauma and inflammation, we also tested traits related to behavioral and emotional response to trauma, exposure to traumatic events, and the presence of social support. Additionally, since socioeconomic status has a well-established relationship with the traits investigated (i.e., PTSD, CRP, traumatic events, and social support),^20-22^ we tested whether the associations observed are independent from the effects that are mediated by socioeconomic status (SES).

## Materials and methods

### Cohorts Investigated

Genome-wide association data for PTSD were obtained from PGC-PTSD using Freezes 1.5 and 2. PGC-PTSD Freeze 2 includes 23,185 cases and 151,309 controls of European descent, including both PGC and UKB samples. PGC-PTSD Freeze 1.5 includes only 12,823 cases and 35,648 controls available from PGC samples (i.e., UKB samples were not included in the Freeze 1.5 meta-analysis). We used the PGC-PTSD Freeze 1.5 dataset to exclude the potential bias induced by the overlap between PGC-PTSD Freeze 2 and UKB samples in the PRS and MR analyses. PTSD diagnosis across PGC-PTSD cohorts was obtained considering different version of DSM criteria (DSM-III-R, DSM IV, DSM-5). A detailed description of the QC criteria and GWAS methods was reported previously.^23^

To investigate inflammatory processes, we used the genome-wide data generated by the CHARGE Inflammation Working Group, which investigated the genetics of circulatory CRP levels measured via standard laboratory techniques in up to 148,164 subjects of European ancestry.^24^

Additionally, we obtained GWAS summary association data related to trauma traits, behavioral and emotional response to trauma, exposure to traumatic events, and the presence of social support from the UK Biobank (UKB)^25^ included in the category “traumatic events” (UKB Field ID: 145). These were assessed via the UKB online mental health questionnaire (up to 118,000 individuals of European descent).^26^ In addition, we investigated indicators of SES, considering “Average total household income before tax” (UKB Field ID: 738, household income) and “Townsend deprivation index at recruitment” (UKB Field ID: 189, deprivation index). ^27^ Details regarding QC criteria and GWAS methods of this previous analysis are available at https://github.com/Nealelab/UK_Biobank_GWAS/tree/master/imputed-v2-gwas. Briefly, the association analyses for all phenotypes were conducted using regression models available in Hail (available at https://github.com/hail-is/hail) including the first 20 ancestry principal components, sex, age, age^2^, sex×age, and sex×age^2^ as covariates. Details regarding the UKB traits investigated in this study are reported in Supplemental Table 1.

**Table 1:** Significant PRS results for CRP, PTSD, trauma and socioeconomic traits.

| Base | Target | R<sup>2</sup> | FDR Q-value |
| --- | --- | --- | --- |
| Household income | CRP | 0.08% | 7.45E-25 |
| Household income | PTSD | 0.01% | 1.36E-07 |
| Deprivation index | PTSD | 0.01% | 6.00E-05 |
| CRP | Physically abused by family as a child | 0.01% | 1.80E-04 |
| Deprivation index | CRP | 0.01% | 2.70E-04 |
| PTSD | Household income | 0.002% | 5.52E-04 |
| CRP | Able to pay rent/mortgage as an adult | 0.010% | 8.10E-04 |
| CRP | Felt loved as a child | 0.012% | 4.20E-03 |
| Felt loved as a child | CRP | 0.01% | 1.60E-02 |
| PTSD | CRP | 0.01% | 2.70E-02 |
| CRP | PTSD | 0.01% | 3.70E-02 |
| CRP | Household income | <0.001% | 3.70E-02 |

### Heritability and Genetic Correlation

To estimate observed-scale SNP-heritability and genetic correlation among the phenotypes interrogated in this project (PGC-PTSD Freeze 2, CRP, trauma traits, and socioeconomic status traits), we performed Linkage Disequilibrium Score Regression (LDSC) according to the pipeline described previously (available at https://github.com/bulik/ldsc).^28^

We also calculated the heritability z-score, which is defined as the heritability estimate produced by LDSC divided by its standard error. The heritability z-score is able to capture information about the genetic architecture of a trait and the reliability of each heritability estimate. As recommended by LDSC best practice, we conducted genetic correlation analysis only using traits with heritability z-score>4.^28^ Genetic correlations were adjusted for multiple testing using false discovery rate (FDR), considering Q < 0.05 as the significance threshold.

### Polygenic Risk Score (PRS) and Definition of the Genetic Instruments

PRS were calculated using PRSice v1.25 software (available at http://prsice.info/)^29^ and considering the following parameters: clumping with a linkage disequilibrium (LD) cutoff of R^2^ = 0.001, 10,000-kb as clumping window, and exclusion of major histocompatibility complex (MHC) region of the genome because of its complex LD structure. European samples from the 1000 Genomes Project were used as the LD reference panel.

To conduct this PRS analysis, we used GWAS summary data from the cohorts described above (PGC-PTSD freeze 1.5 - to avoid sample overlap between PGC-PTSD Freeze 2.0 and UKB). We applied FDR multiple testing correction to correct the p-value for the number of PRS p-value thresholds tested at Q < 0.05.

### Mendelian Randomization (MR)

We used the R package TwoSampleMR^30^ to infer the causal relationship between the phenotypes of interest, testing genetic variants (i.e., SNPs) which survived the PRS FDR correction as instrumental variables. This R package permits conducting five MR analysis: inverse variance weighted (IVW), MR-Egger, weighted median, simple mode, and weighted mode.^30^ These methods were considered to verify the stability of the causal effects of MR results and the IVW approach was reported as the main results due to its higher statistical power.^31^ The MR analysis was conducted in accordance with the STROBE (STrenghtening the Reporting of Observational studies in Epidemiology) - MR reporting guidelines.^32^

Multiple MR sensitivity tests were performed to investigate the presence of horizontal pleiotropy and heterogeneity among the variants included within the genetic instruments tested: MR-Egger regression intercept,^33^ MR-PRESSO (Pleiotropy RESidual Sum and Outlier) global test),^34^ and MR heterogeneity test.^35^ Outliers contributing to the heterogeneity and horizontal pleiotropy within the genetic instruments were identified via a leave-one-out analysis and considering MR-RAPS (MR–Robust Adjusted Profile Score)^36^ standardized residuals that fall outside the 95% confidence level.

Multivariable MR (MVMR) was applied to estimate the effect of two or more traits (risk factors) on a specific outcome (disease) using the multivariable IVW approach available in the MendelianRandomization R package.^37^

## Results

### Genetic Correlation and Polygenic Risk Scores analyses

SNP-heritability was estimated for all traits investigated in this study and we found that 4 trauma response traits, 12 trauma exposure traits, and 2 socioeconomic status traits had heritability z-score>4 (Supplemental Table 2). We found a positive genetic correlation between CRP and PTSD diagnosis (PGC-PTSD Freeze 2: rg=0.155, p=0.026). After multiple testing correction (FDR at 5%), we observed significant genetic correlation of CRP with seven traits related to behavioral and emotional response to trauma, exposure to traumatic events, and the presence of social support (Figure 1). Positive genetic correlation was also observed between CRP and socioeconomic status: deprivation index, rg=0.20, p=3.38×10^−5^; household income, rg=-0.25, p=4.08×10^−10^. The direction of the genetic correlations of CRP was in line with expectations: positive genetic correlation with the presence of PTSD, traumatic events, and deprivation index; negative genetic correlation with social support and household income.

**Figure 1:**
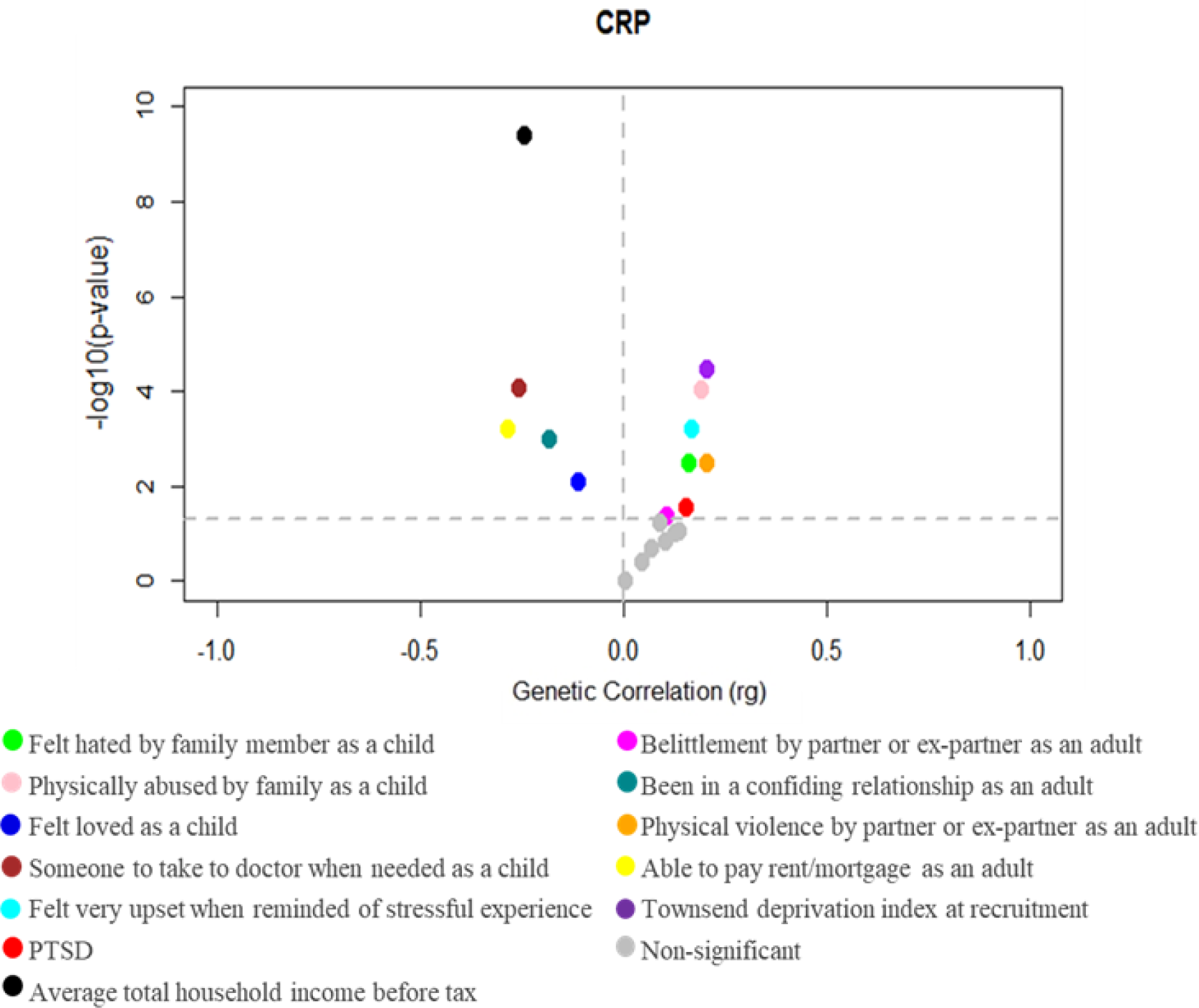
Genetic correlation of CRP with PTSD, behavioral and emotional response to trauma, exposure to traumatic events, and the presence of social support.

Considering our genetic correlation findings, we conducted bidirectional PRS between CRP and trauma traits, PGC-PTSD Freeze 1.5 case-control status, and socioeconomic traits. After FDR 5% correction for the number of PRS thresholds, we found 12 significant associations (Table 1). The most significant PRS association was observed between household income and CRP (R^2^=0.08%; FDR Q-value=7.45×10^−25^).

### Mendelian randomization (MR)

Based on the PRS findings, we performed two-sample MR to assess the causality among the traits. If heterogeneity or pleiotropy was identified within the genetic instruments, we removed the outlier variants to verify that the causal estimates were not generated by potential biases within the genetic instruments.

In our MR analysis, we found evidence supporting several causal relationships among the traits investigated (Figure 2; Supplemental Table 3). Our results suggest a bidirectional association between PTSD and CRP, where genetically determined CRP levels have a causal effect on PTSD (β=0.065, 95% CI: 0.012 to 0.119, p=0.015), and genetic liability to PTSD has a causal effect on CRP (β=0.008, 95% CI: 0.001 to 0.015, p=0.024). We observed the presence of possible heterogeneity in SNPs evaluated in the PTSD→CRP association (heterogeneity test p=0.001). However, after removal of the outlier variants from the genetic instrument, the PTSD→CRP association remained significant (β=0.008, 95% CI: 0.002 to 0.015, p=0.009).

**Figure 2:**
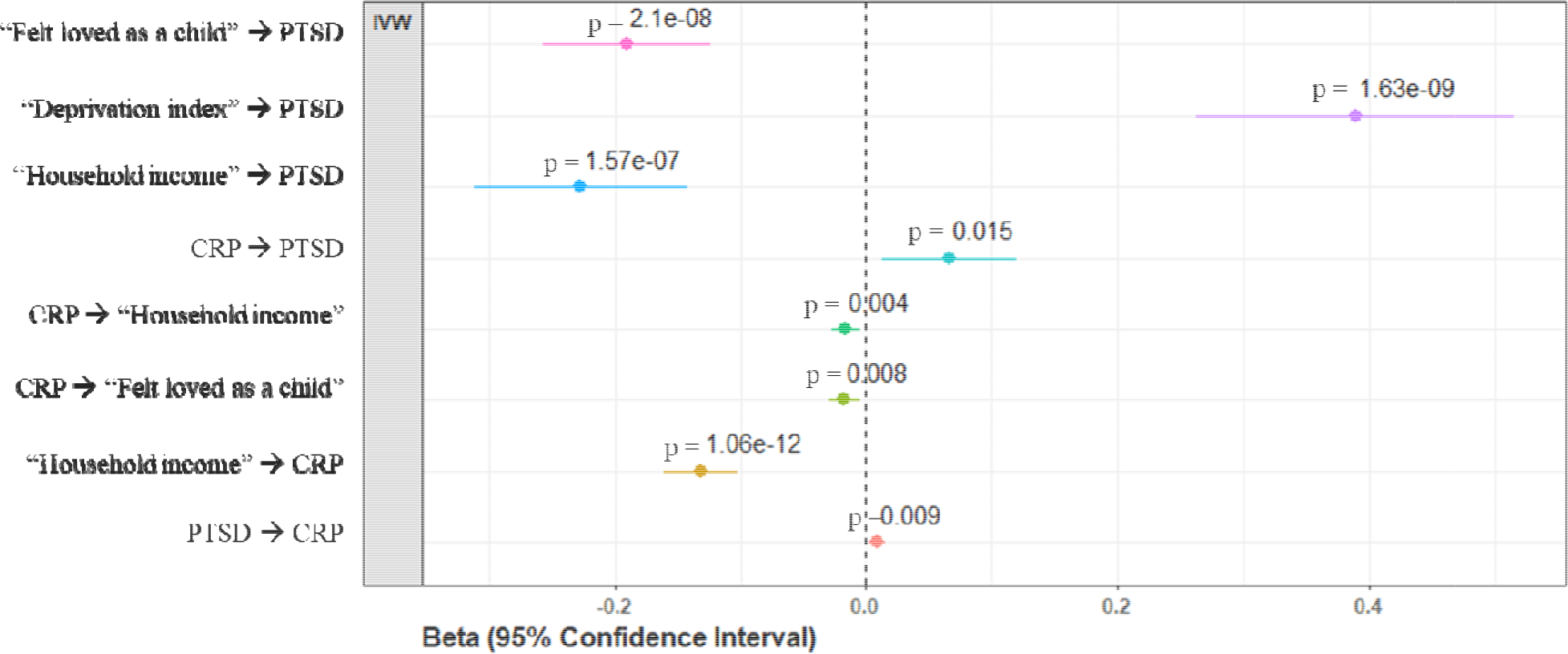
Significant two-sample MR results.

Among the traits related to traumatic events and social support, we observed that CRP also has a negative causal effect on the “Felt loved as a child” trait from UKB (β=-0.02, 95% CI: −0.034 to −0.007, p=0.003). However, strong evidence of heterogeneity was present among the variants included in the genetic instrument (heterogeneity test p=4.26×10^−5^). Removing the variants contributing to heterogeneity within the genetic instrument, we confirmed the CRP-“Felt loved as a child” association (β=-0.017, 95% CI: −0.031 to −0.004, p=0.008). “Felt loved as a child” trait showed a protective effect on PTSD risk (β=-0.139, 95% CI: −0.206 to −0.071, p=5.64×10^−6^). However, the genetic instrument was affected by heterogeneity (heterogeneity test p 0.002). Removing the variants generating the heterogeneity within the genetic instrument, we confirmed the protective effect of “Felt loved as a child” trait on PTSD (β=-0.19, 95%CI=-0.124 to - 0.257, p=2.1×10^−8^).

We also observed that indicators of SES affect both CRP levels and PTSD risk. Household income showed a negative bidirectional association with CRP (household income→CRP β=-0.131, 95%CI=-0.161 to −0.102, p=1.06×10^−18^; CRP→household income β=-0.016, 95%CI=-0.027 to −0.005, p=0.004). Both household income and deprivation index have a causal effect on PTSD (Household income: β=-0.277, 95% CI: −0.312 to −0.142, p=1.57×10^−7^; Deprivation index: β=0.389, 95% CI: 0.262 to 0.515, p=1.63×10^−9^). The effects were confirmed also after removing outlier variants when heterogeneity or horizontal pleiotropy was observed within the genetic instruments (Supplemental Table 3)

Finally, we performed a MVMR analysis to verify whether the causal effects observed among PTSD, CRP, social support, and SES are independent from each other (Figure 3). We confirmed that genetically determined CRP has causal effect on PTSD, independently of the “Felt loved as a child” association (β=0.066, 95% CI: 0.007 to 0.126, p=0.029; Figure 3A). However, the CRP→PTSD association (univariate MR β=0.066, 95% CI=0.013 – 0.119, p=0.015; multivariate MR β=0.043, 95% CI=-0.027 – 0.113, p=0.232) was not independent from the effect of deprivation index (Deprivation index→PTSD: univariate MR: β=0.38, p=1.63×10^−9^, 95% CI=0.263 – 0.516; multivariate MR β=0.27, p=0.016; 95% CI= 0.050 – 0.498; Figure 3B) and household income (Household income→PTSD: univariate MR β=-0.22, 95% CI=-0.312 – −0.142, p=1.57×10^−7^; multivariate MR β=-0.17, 95% CI=-0.294 – 0.051, p=0.005; Figure 3C).

**Figure 3:**
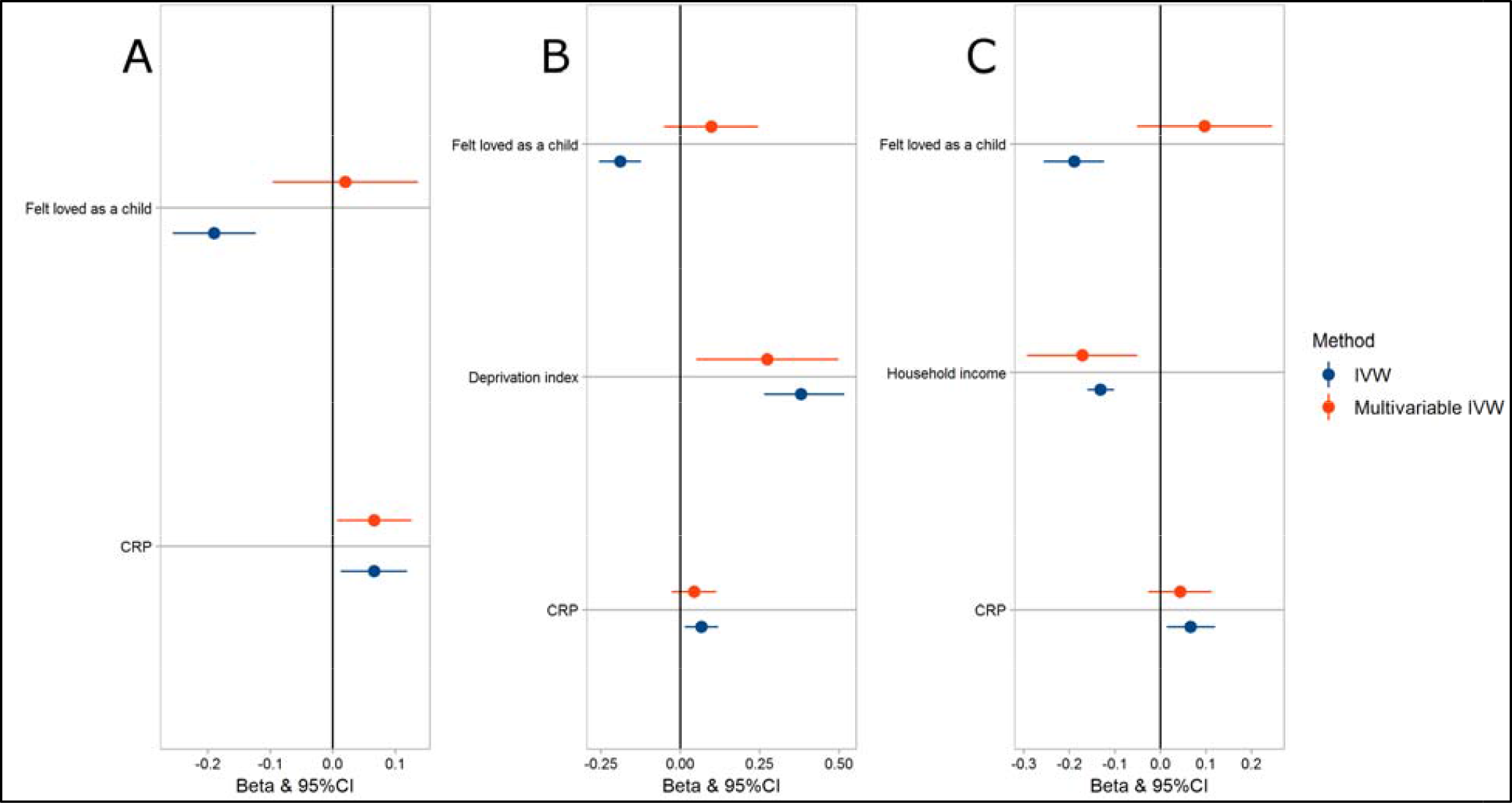
Multivariable MR results considering the following models: **A**. Felt loved as a child + CRP → PTSD; **B**. Felt loved as a child + Deprivation index + CRP → PTSD; **C**. Felt loved as a child + Household income + CRP → PTSD.

## Discussion

We used genome-wide data to investigate the genetic overlap and the causal mechanisms between CRP levels and PTSD, also considering the effects of SES, traumatic experiences, and social support. Our findings indicate that there is bidirectional association between PTSD and CRP levels, which appears to be strongly affected by SES.

CRP is a protein that responds to inflammatory stimuli by triggering cellular reactions and it is normally induced through pro-inflammatory cytokines, mainly by interleukin-6 secretion.^38, 39^ CRP may be produced in the microvessel endothelial cells that form the blood-brain-barrier, and peripheral CRP can affect the central nervous system via mechanisms of blood-brain barrier disruption associated with elevated levels of peripheral inflammatory cytokines.^40, 41^ Higher levels of pro-inflammatory cytokines may influence neurotransmission, leading to altered production of serotonin, norepinephrine, dopamine and brain-derived neurotrophic factor. This effect on levels of neurotransmitters may be associated with specific psychiatric symptoms, such as anhedonia.^42^ Furthermore, these cytokines may influence the neurocircuitry, causing alterations in motivation status, and anxiety, arousal, and alarm response.^43^ Twin and family studies estimated that heritability of CRP is around 35%-40%.^24^ Several environmental factors, such as sociodemographic variables, dietary habits, behavioral and lifestyle factors, are also strongly associated with CRP level.^44^ Previous cross-sectional and longitudinal studies highlighted the presence of a bidirectional association between PTSD and CRP levels.^6-13^ Our findings confirm this scenario. However, we also observed that the CRP-PTSD association is not independent from SES (i.e. high deprivation index and low household income). The effect of SES on CRP levels was previously observed in studies highlighting the association of increased CRP levels with low SES and traumatic childhood experiences.^45^ In addition, low SES has been associated with DNA methylation of genes involved in inflammation, suggesting that alterations in epigenetics of specific genes might influence the inflammation response leading to increased inflammatory markers.^46, 47^

A recent GWAS of social stratification in Great Britain showed that genetic factors related to socio-economic status (educational, income, employment) are associated with a wide range of physical and mental health outcomes.^48^ Lower economic status might be a mediator of the negative association between educational attainment and PTSD.^49^ In line with these previous findings, we found that SES might affect the interplay among inflammatory processes (i.e., CRP level), PTSD, and childhood social support. This confirms that SES may affects several of the comorbid traits associated with PTSD.

Furthermore, we found that “Felt loved as a child” was a protective factor with respect to genetic liability to PTSD and CRP levels. Social support systems have been considered as an important protective factor for mental health and well-being of individuals, and refer to social interaction which involves love, care, respect, affection, acceptance, and financial assistance.^50, 51^ Social support might influence trauma perception and cognitive processes and consequently reduce the risk to develop psychiatric disorders. Individuals with low social support had worse prognosis of PTSD.^52, 53^ Additionally, high levels of social support were associated with reduced CRP levels.^54, 55^

Although we present novel results contributing to understand the association among PTSD, CRP levels, traumatic events, and social support, our study has several limitations. Our analyses focused on CRP levels only, because no large-scale GWAS has investigated other inflammatory biomarkers. Further studies will be needed to verify whether the same effects are present across inflammation-related molecules such as IL-6. The analyses were conducted using GWAS in subjects of European ancestry, thus we cannot generalize these results to other populations. The magnitude of estimates for the effect of CRP on PTSD is relatively small, which may indicate a minor contribution of the associations reported in the biology of PTSD. The genetic information about PTSD and trauma traits were obtained across different cohorts, which may cause heterogeneity in the outcomes related to stress exposition. The datasets investigated were generated from participants assessed using different instruments, also including online questionnaires. Although this approach was needed to collect information from a large cohort and the online instruments were validated,^26^ this may have affected the quality of the data collected.

In conclusion, this study is the first analysis leveraging large-scale genome-wide datasets to investigate the underlying mechanisms linking CRP, PTSD, and traumatic experiences. Our findings support a bidirectional association between PTSD and CRP, which appears to be strongly affected by SES. Additionally, SES traits seem to also mediate the interplay among CRP levels, PTSD, and childhood emotional support.

## Data Availability

All data generated during this study are included in this published article and its Supplemental information files.

## Acknowledgements

This research was supported by the Veterans Affairs National Center for Posttraumatic Stress Disorder Research, the Simons Foundation Autism Research Initiative (SFARI Explorer Award: 534858) and the American Foundation for Suicide Prevention (YIG-1-109-16), and R01MH106595. C.M.C. and S.I.B. were supported by Fundação de Amparo à Pesquisa do Estado de São Paulo (FAPESP 2018/05995-4) international fellowship and Coordenação de Aperfeiçoamento de Pessoal de Nível Superior (CAPES Code 001).

